# Compositional microbiome-based signatures associate with general health status: findings from a large population-based cohort study

**DOI:** 10.64898/2026.06.03.26354796

**Authors:** M. Pujolassos, A. Kurilshikov, R.K Weersma, J. Fu, A. Zhernakova, M.L. Calle

**Affiliations:** Department of Bioscience, University of Vic – Central University of Catalonia, Vic, 08500, Spain; Department of Genetics, University of Groningen, University Medical Center Groningen, Groningen, 9713 GZ, The Netherlands; Department of Gastroenterology and Hepatology, University of Groningen, University Medical Center Groningen, Groningen, 9713 GZ, The Netherlands; Department of Pediatrics, University of Groningen, University Medical Center Groningen, Groningen, 9713 GZ, The Netherlands; Institut de Recerca i Innovació en Ciències de la Vida i de la Salut a la Catalunya Central (IRIS-CC), Vic, 08500, Spain

**Keywords:** compositional data analysis, population-based cohort, microbial signatures, general health

## Abstract

While microbiome is increasingly recognized as crucial for human health, translating this knowledge into effective healthcare and preventive strategies remains challenging. Many studies focus on identifying changes in microbiome composition associated with disease and evaluating the potential of such disease-associated microbial profiles as biomarkers for disease diagnosis. Under the hypothesis that microbiome dysbiosis may reflect physiological alterations present long before disease onset, in this work, we analyse the potential of disease-specific microbial signatures not as a diagnostic tool when the disease is already present, but as a means of health assessment in the general population. Moreover, instead of trying to define a single health measure, we believe it is necessary to consider several ways in which the microbiome departs from health, according to different disease-related physiological changes.

To evaluate our assumptions, we designed a two-stage study: the identification of disease-specific microbial signatures (discovery stage) and, subsequently, the study of their distribution in the general population to assess associations with general health (external validation stage). Specifically, in the discovery phase we characterized 16 disease-specific bacterial signatures from large public microbiome data using a compositional data analysis methodology. In the second phase, we quantified these microbial signatures in the Lifelines-DMP cohort, a large population-based cohort, and evaluated their association with self-reported health status.

Results indicate that most disease-specific microbial signatures associate with health status, supporting our assumption that microbial composition can capture physiological alterations before disease onset, and highlighting the importance of considering multiple ways in which microbiome departs from a healthy state. These findings reaffirm the potential of microbial information as an additional tool in preventive medicine.

## Introduction

For certain infectious diseases, the presence of specific pathogenic bacteria is the direct cause of disease, as described by Koch’s postulates [1]. In contrast, for most diseases, microbial associations are characterized by shifts in the overall microbiome composition, involving simultaneous over- and under-representation of multiple bacterial species. Identifying these imbalanced ecologies, which deviate from the “healthy” balanced state, is the basis for constructing predictive microbiome models of disease. The set of microbes that constitute such imbalances, also called microbial signatures, could be a potential tool for disease detection.

Translating and applying the knowledge of human microbiome into healthcare promotion remains challenging. Some attempts have been done for specific diseases like atopic dermatitis [2], stroke [3], bacterial vaginosis [4] or colorectal cancer [5], among others. However, the characterization of microbial signatures for health prediction is constrained by important challenges concerning large inter-individual heterogeneity, high technical variations, and overall small sample sizes. These are some of the reasons why results are usually inconsistent between microbiome studies and thus having little reproducibility. Despite efforts of large population-scale microbiome projects to describe human microbiome [6–8], it hinders a consensus definition of a healthy and a diseased microbiome. In 2020, Gupta and collaborators proposed a gut microbiome index to predict health status, *i*.*e*., presence or absence of disease [9]. They analysed 4347 human stool samples across different studies and health conditions and defined two groups of bacteria based on their prevalence in healthy compared to non-healthy subjects. They identified 7 “health-prevalent” bacteria, more frequently observed in healthy subjects and 43 “health-scarce” bacteria, less frequent in healthy individuals. Their Gut Microbial Health Index, GMHI, is expressed as a particular ratio between health prevalent and health-scarce bacterial groups. GHMI achieved high reproducibility in the classification of healthy and non-healthy individuals in independent cohorts, with classification accuracies that ranged from 70.1% for healthy subjects to 77.1% for non-healthy subjects [9]. The robustness of the method was further corroborated by Gacesa [10] when GMHI was replicated into a Dutch general population cohort and resulted in significant differences between healthy individuals compared to subjects suffering from any disease. Moreover, the Dutch microbial signature associated with health replicated 43 out of 50 features comprising GHMI, suggesting the existence of a common bacterial signature across diseases that characterizes diseased from healthy individuals.

Instead of searching for a univariate health index for distinguishing between healthy and diseased gut microbiomes, in the present work we consider multiple ways of departing from a healthy state, according to specific physiological processes altered during disease. Under the hypothesis that microbiome dysbiosis may reflect physiological alterations present long before disease onset, in this work, we analyse the potential of disease-specific microbial signatures not as a diagnostic tool when the disease is already present, but as a means of health assessment in the general population. Our approach for testing this hypothesis consists of two stages: the identification of disease-specific microbial signatures (discovery stage) and, subsequently, the study of their distribution in the general population to assess associations with general health (external validation stage). Under a compositional data analysis (CoDA) framework, in the first discovery step, we characterized disease-specific microbial signatures from a large curated public microbiome data [11] using *coda4microbiome* methodology [12]. These signatures reflect different patterns of microbiome composition that differ from cases and controls. The second step consisted of an assessment of the disease-specific microbial signatures capacity for correlating with general health status on an external population cohort, the Lifelines DMP [10]. The cohort comprises metagenomic, clinical and anthropomorphic information of 8,208 individuals from the general population of the northern Netherlands. We quantified the disease-specific signatures in the general population cohort and evaluated their association with the self-reported health status of the individuals. Finally, we also explored the potential of microbiome to predict general health and well-being in the general population.

## Materials and Methods

### Identification of disease-specific microbial signatures from a public repository

A compositional data analysis approach, *coda4microbiome* [12], was used to characterise disease-specific bacterial signatures from faecal microbiome samples in the large public repository curatedMetagenomicData [11]. Bacterial signatures provided by *coda4microbiome* account for the compositionality of microbiome data and are expressed as a log-contrast function, *i*.*e*., a linear combination of log-transformed abundances with the sum of the coefficients being zero. This ensures the scale invariance principle of CoDA and provides an interpretation of the signature as a weighted balance between two groups of bacteria: some features contributing positively to the outcome and others negatively. After exploring 20 different diseases, we obtained a total of 16 disease-specific bacterial signatures with enough association with the disease to be worth considering for their external validation on the general population cohort Lifelines-DMP: signatures for cirrhosis, CAD (coronary artery disease), HF (heart failure), IGT (impaired glucose tolerance), T2D (type 2 diabetes), ACVD (atherosclerotic cardiovascular disease), STH (soil transmitted Helminths), ME/CFS (myalgic encephalomyelitis/chronic fatigue syndrome), CRC (colorectal cancer), adenoma, BD (Bechet’s disease), PD (Parkinson disease), schizophrenia, IBD (inflammatory bowel disease), IBD-UC (ulcerative colitis) and IBD-CD (Crohn’s disease). For more detailed information about the discovery stage of the disease-specific signatures see Supplementary Information (**Fig. S1**).

### Quantification of disease-specific bacterial signatures in a population-based cohort

The Lifelines-DMP cohort [10] is a general population cohort from the northern Netherlands that comprises stool metagenomic information of 8,208 volunteered individuals. Detailed self-reported information about participant’s health status and medication use was compiled from questionnaires and additional measurements were collected from participant’s blood and urine sample. Participants ranked their general health as excellent, very good, good, mediocre, or poor, representing their self-perceived well-being. In this study, general health was analysed in three ways: as an ordinal numeric variable (GH), a binary variable (GHbi) grouping “good health” (excellent to good) vs. “bad health” (mediocre and poor), and another binary version (GHbix) including only “excellent” in the good health group. To minimize microbiome disturbances, participants using antibiotics or PPIs were excluded, resulting in 5,984 samples with information on self-reported general health. This final cohort was 58.7% female, with a mean age of 50.7 years (±11.7) and a mean BMI of 25.8 (±3.9).

Since our disease-specific signatures are combinations of log-transformed bacterial abundances, their quantification in the Lifelines-DMP participants was performed by combining the (log) relative abundances of selected bacterial species with the corresponding model coefficients. This provided a microbial risk score, *M*, for each health condition and every individual in the Lifelines-DMP cohort. All models were specified so that higher scores corresponded to a higher risk of the disease. To avoid indeterminacy associated with the logarithmic transformation of zero values, we replaced zeros by a small quantity (*i*.*e*. half of the minimum relative abundance observed across all abundance values). To construct coherent signatures and avoid spurious results due to zero inflated data, we removed samples with all absent species for a given signature. For this reason, sample sizes differ according to the specific bacterial signature analysed. We then assessed the association between bacterial scores with general health status, as well as their predictive power for healthy phenotype assessment.

### Disease-specific bacterial signatures and general health

In first place, we performed an exploratory analysis of disease-specific bacterial scores across all five GH categories. Moreover, since basal characteristic like sex, age, BMI and diet play important roles in either health and microbiome composition, we explored their role with respect to general health status and also on disease-specific bacterial scores. Taking diet into account becomes a real challenge because dietary patterns are not always recorded in microbiome studies, and when they are, different variables may be recorded. Gut microbial composition of populations from different geographic regions and dietary habits revelled a significant relationship between *Prevotella*/*Bacteroides* (P/B) and diet, making this ratio useful as a tool to reduce variability when specific dietary information is not available [13–15]. Statistical significance of exploratory analyses was carried out by Kruskal-Wallis and Wilcoxon rank-sum tests for numeric variables, and Chi-squared tests for categorical variables. Kendall test was performed to assess trends between general health status and microbial signature scores.

Secondly, we considered regression models that allow adjusting for possible confounders (sex, age, BMI, P/B ratio) and other covariates. We used *glm()* and *lm()* functions from *stats* R package [16] to analyse GH and microbial signatures adjusting for covariates.

In third place, we evaluated the impact of adding microbiome information in predicting general health. We considered two different ways of including microbiome information in the model: α-diversity, measured using the chao1 index, and disease-specific bacterial signatures. Hence, we performed three different regression models: models including basal variables only (sex + age + BMI + P/B), adding α-diversity (sex + age + BMI + P/B + α-diversity), and models comprising the previous variables together with disease-specific microbial scores, *M*, (sex + age + BMI + P/B + α-diversity + M). Prediction accuracy of all regression models was evaluated by AUC and the different models were compared using analysis of variance (ANOVA) for nested models [17].

Finally, we identified associations between bacterial signatures and other health factors like specific diseases, symptoms, or blood measurements. With this purpose, we stratified the population into two groups according to the 90^th^ percentile of the microbial signature score and checked, using the hypergeometric test, whether diseased phenotypes were over-represented, or enriched, in the group of subjects with higher microbial scores (above the 90^th^ percentile). For those numerical health measurements, such as immunological parameters, we performed regression analysis between microbial scores and those measurements with adjustment for basal characteristics. All analyses were conducted with R 4.5.3.

## Results

### Interplay between bacterial signatures, general health and basal characteristics

#### Basal variables contribute to general health phenotype and microbiome

The exploratory analyses of the basal variables (*e*.*g*., sex, age, BMI and *Prevotella/Bacteroides* ratio) confirmed their association to both health and microbiome and corroborated the need to consider them as possible confounders.

General health levels, from excellent to poor, presented gradual differences regarding some of the basal characteristics, especially BMI, P/B and sex (**Fig. 1**). Individuals with worse general health tended to have higher BMI and lower ratio of P/B (Kruskal-Wallis test significance < 0.0001 for BMI and 0.0075 for P/B). Linear regressions confirmed the association of poorer general health with high BMI values and low P/B ratio (P-values for BMI and P/B coefficients of 2.93 x10^-43^ and 9 x10^-4^, respectively). Women reported significant worse general health than men (Chi-squared P-value = 6.4 x10^-7^ and male negative linear regression coefficient significance of 4.29 x 10^-10^). Age instead was not clearly associated with the general health in this cohort (regression coefficient P-value = 0.14). On the other hand, microbial α-diversity of these individuals differed significantly. Subjects with worse general health presented a less diverse microbiome compared to those with better general health (chao1 Kruskal-Wallis P-value < 0.001 and negative linear regression coefficient significance of 1.23 x 10^-9^). Similar results were obtained for general health when considered a binary outcome variable (GHbi and GHbix) (**Fig. S2**).

**Fig. 1.**
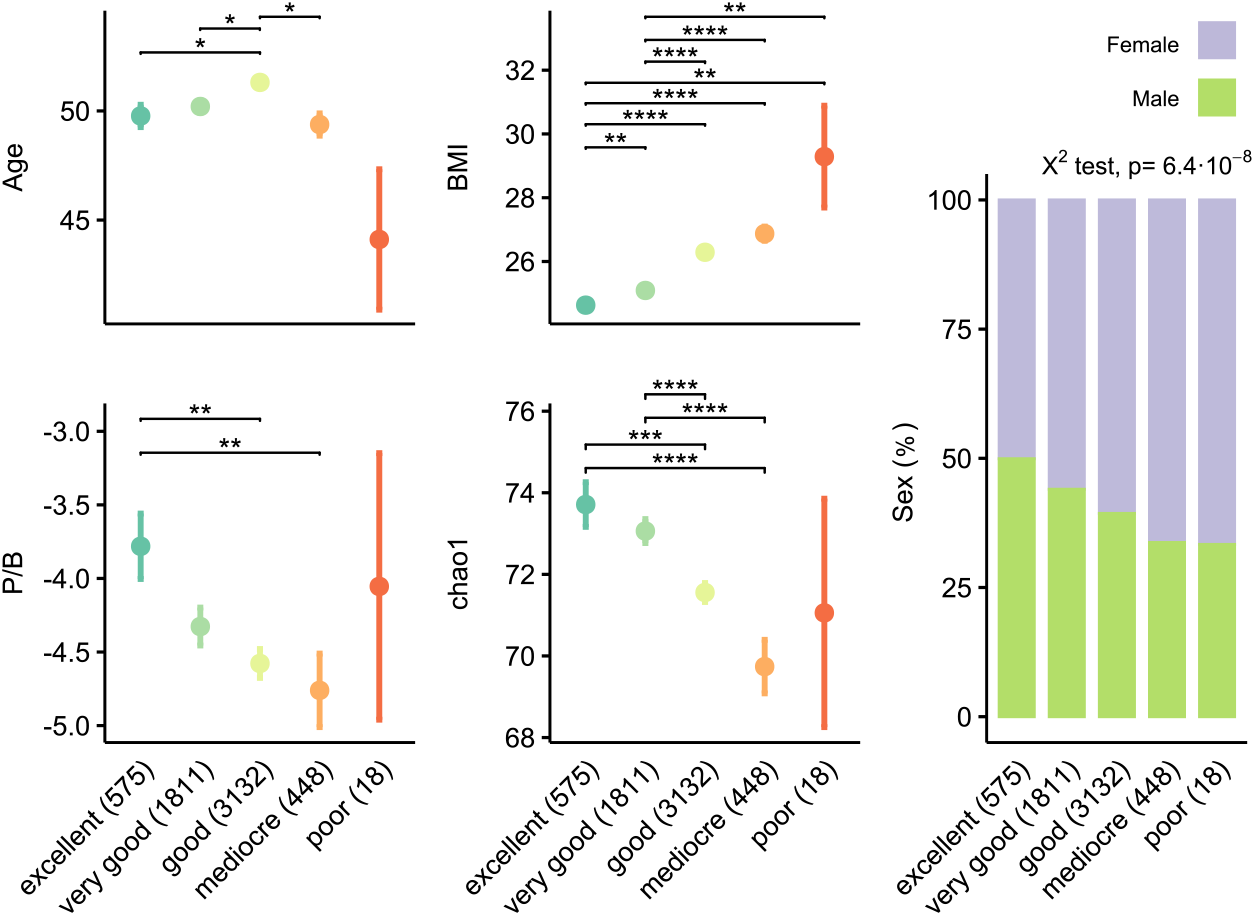
Basal variables across General Health categories. Mean and confidence intervals for age, BMI, P/B, chao1 and sex are represented (vertical axis) for all five categories of GHnum, including “excellent”, “very good”, “good”, “mediocre” and “poor” (horizontal axis). The number of samples within every group is reported in brackets (n). Wilcoxon adjusted P-value significance: **** ≤ 0.0001, *** ≤ 0.001, ** ≤ 0.01, * ≤ 0.05

We also explored the relationship between basal variables and all 16 microbial signatures. We identified significant sex differences in microbial scores with females having higher scores than males for most of the signatures. Chao1 index presented significant negative correlations with all bacterial signature scores. The remaining covariates, age, BMI, and P/B did not show important correlations with bacterial signatures (data not shown).

#### Disease-specific bacterial signatures associate with General Health

We were particularly interested in characterizing disease-specific bacterial signatures in the general population and assess if larger signature scores could be indicative of certain level of health worsening. These analyses identified positive associations between signatures scores and general health for 11 (out of 16) bacterial signatures, 9 of them showed significant (**Fig. 2**). Individuals with worse general health phenotypes presented significant higher values of microbial scores for adenoma, CAD, cirrhosis, CRC, HF, IBD-UC, IBD, STH and T2D signatures compared to groups with better general health. Similar results were obtained for IGT and IBD-CD, though the results were not significant. Significant positive Kendall’s tau between increasing bacterial scores and poorer general health status confirmed the observed trend of the above mentioned signatures (**Table S3**). Similar results were obtained for these signatures when comparing good *vs*. bad general health phenotypes from GHbi and GHbix (**Fig. S3**). The other 5 signatures (schizophrenia, PD, ME/CFS, BD and ACVD) did not show any pattern related to general health categories (results not shown).

**Fig. 2.**
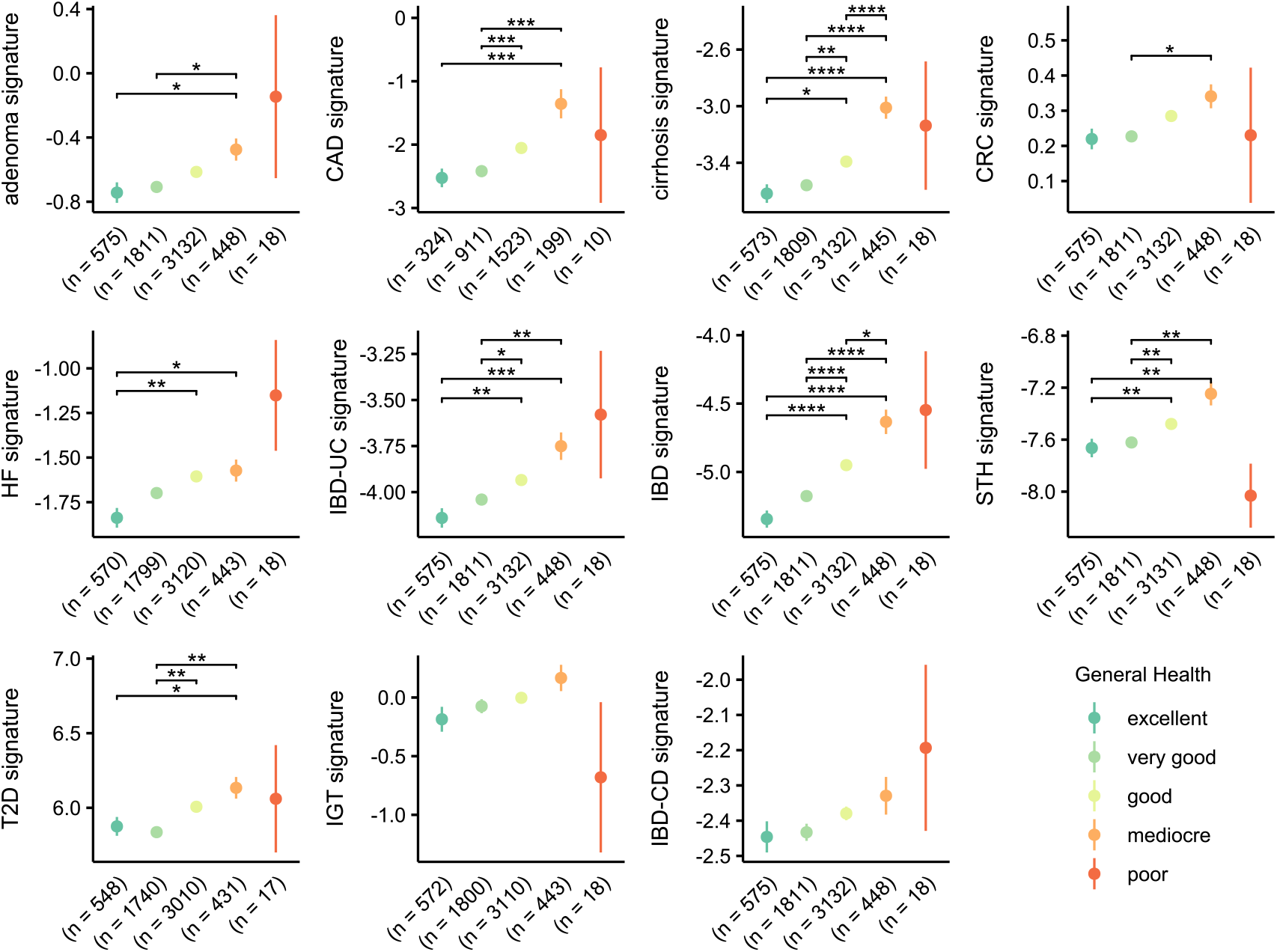
Microbial signature scores of Lifelines-DMP individuals across GH categories. Dots represent mean values of microbial signature score with positive and negative confidence intervals as error bars (vertical axis). All general health categories are represented across horizontal axis and their sample size is represented in brackets (n). Wilcoxon adjusted P-value significance: **** ≤ 0.0001, *** ≤ 0.001, ** ≤ 0.01, * ≤ 0.05. Abbreviations: HF (heart failure), IBD-UC (ulcerative colitis), STH (soil transmitted Helminths), T2D (type 2 diabetes), CAD (coronary artery disease), CRC (colorectal cancer), IBD-CD (Chron’s disease) and IGT (impaired glucose tolerance)

### Regression analysis adjusting for covariates

Given the significant impact of some basal covariates and α-diversity on general health status of the studied cohort, and to avoid cofounding effects, we corrected for sex, age, BMI, P/B and α-diversity when analysing the relationship between general health and bacterial signatures. The adjusted regression models for the different bacterial signatures supported the results found in the exploratory analyses: worse health phenotypes were positively associated with higher microbial scores. Bacterial signatures that maintained their significant association with general health after adjusting for basal variables and α-diversity were IBD, CAD, IBD, IBD-UC, cirrhosis, HF and adenoma. In that line, statistically significant ANOVA for nested models indicated that regression models including these signatures outperformed basal models (models without signature score, *M*). BD resulted to be significantly negatively associated with GH, but not when analysed as binary or extreme binary GH phenotypes. Signatures that did not show any significance in the exploratory analyses (*e*.*g*., schizophrenia, ME/CFS, ACVD, BD), also lacked significance in the regression models (**Fig. 3**). Similar results were found for GHbi and GHbix (**Fig. S4**).

**Fig. 3.**
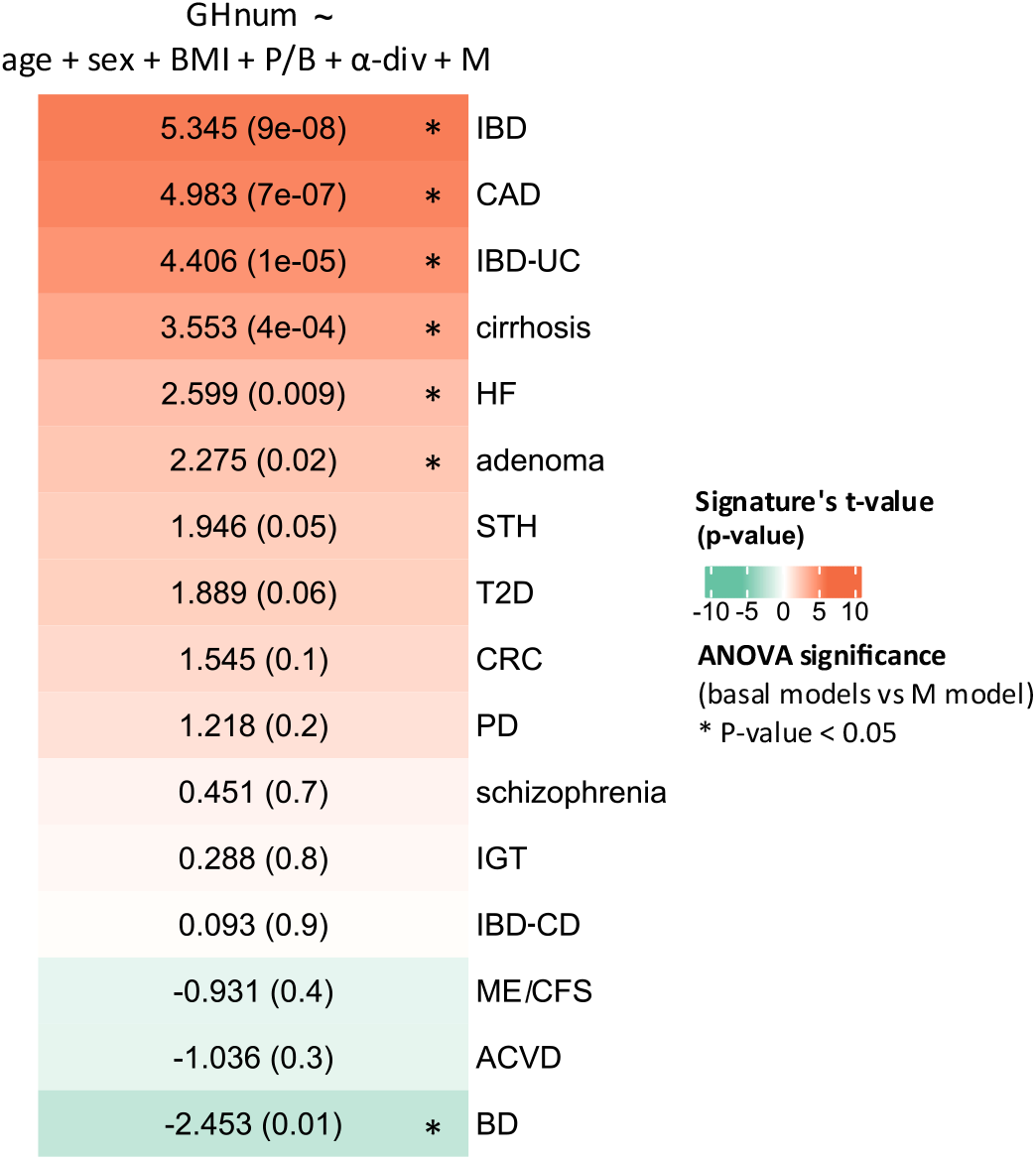
Microbial signatures association with general health. Heatmap colours represent signature’s t-value from every regression model performed with the bacterial signature, M, (their significance reported by P-values in brackets). Positive values are for signature’s association with poorer general health, while negative values indicate association with better general health. M models were compared to basal models with ANOVA for nested-models and its statistical significance is indicated with (*)

### Disease-specific microbial signatures add predictive power to basal models

Since we found positive association between some of the signatures and general health, we tested the additional contribution of microbiome to the prediction of general health status, compared to the basal model including sex, age, BMI and P/B. In this case, we considered two different ways of introducing microbiome composition information: chao1 index, a measure of intra-sample diversity, and the different bacterial signatures.

To assess the prediction capacity of microbial signatures we focused on the binary GH phenotypes (GHbi and GHbix) and compared three models: the basal model, the α-diversity model and the microbial signature model. We then tested the significance of the improvement gained by adding microbial information to the basal model through ANOVA for nested models.

Predicting extreme good *vs*. bad general health (GHbix) performed better than predicting binary GH, although both strategies presented similar results (**Fig. 4** and **Fig S5**). In all cases, adding α-diversity to basal models, improved prediction of GHbix categories, reaching AUC values > 0.7. Furthermore, the addition of microbial signatures slightly enhanced prediction, compared to models including only α-diversity. Models that significantly increased their AUC with the addition disease-specific bacterial signatures were IBD (0.721), IBD-UC (0.713), cirrhosis (0.711) and HF (0.709) (**Fig. 4**). These results were confirmed by the ANOVA for nested models for GH models, in which models including IBD, IBD-UC, CAD, cirrhosis, HF, and adenoma signatures presented significant improvement with respect to basal and α-diversity models (**Table S4**). Behcet’s disease (BD) performed significant improvement when analysing the GH phenotype, but this was not replicated in the binary, or binary extreme GH models.

**Fig. 4.**
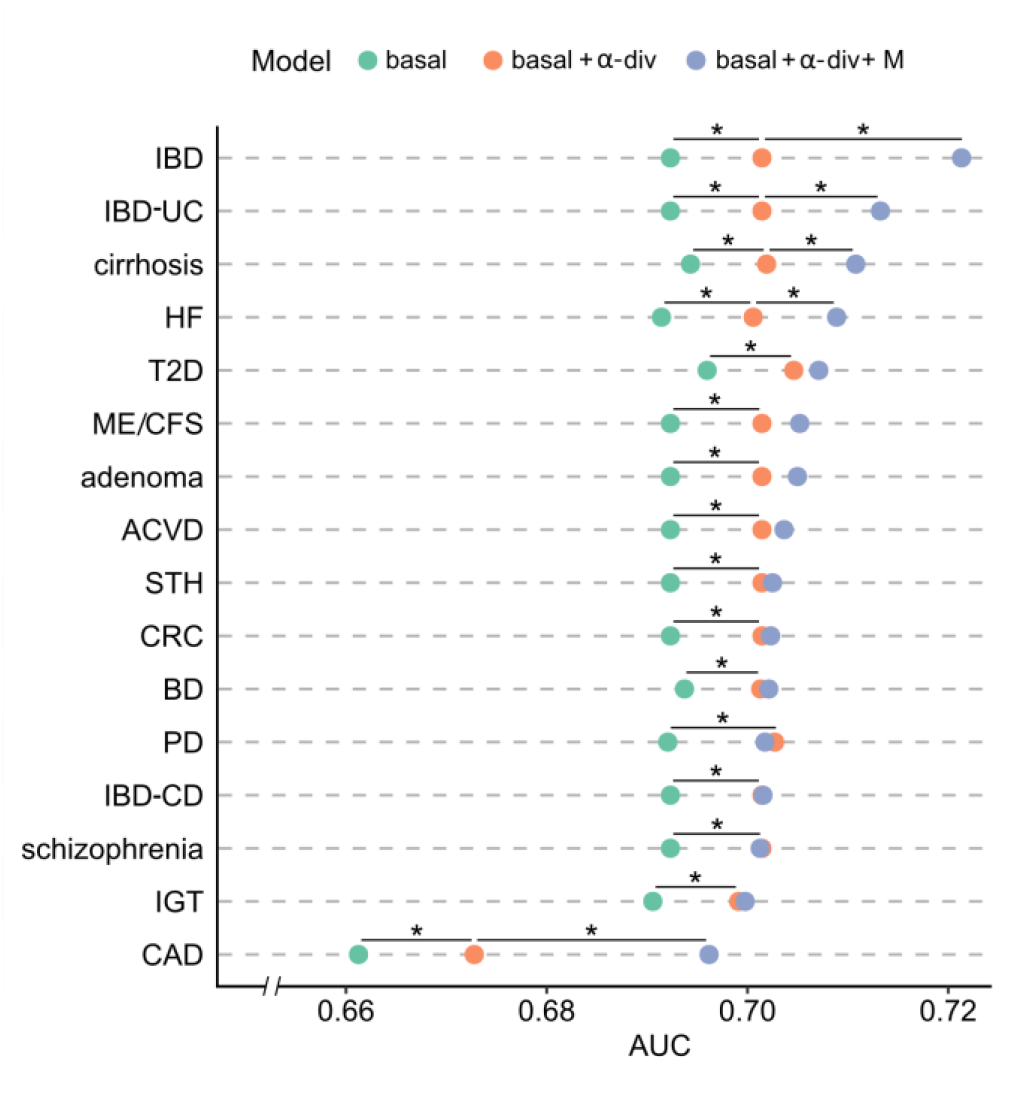
Contribution of microbiome in predicting general health (GHbix). The prediction performance of basal models, models with α-div, and models with α-div + bacterial signatures (M) in terms AUC in the horizontal axis. (*) is for significant improvement between models, assessed with ANOVA for nested models, within 95% confidence level

### Bacterial signatures associate with chronic diseases and other health factors

We explored whether signatures contributing significantly to general health phenotypes might also be associated with other health factors, such as chronic health conditions, symptoms of certain diseases, or specific blood parameters. For each signature, we performed an enrichment analysis comparing comorbidities of individuals with the highest bacterial signatures and those of the rest of individuals in the cohort and identified the 10 chronic diseases overrepresented in the highest risk group. Below, we present a selection of enrichment analyses corresponding to the signatures that presented higher associations with general health (**Fig. 5**).

**Fig. 5.**
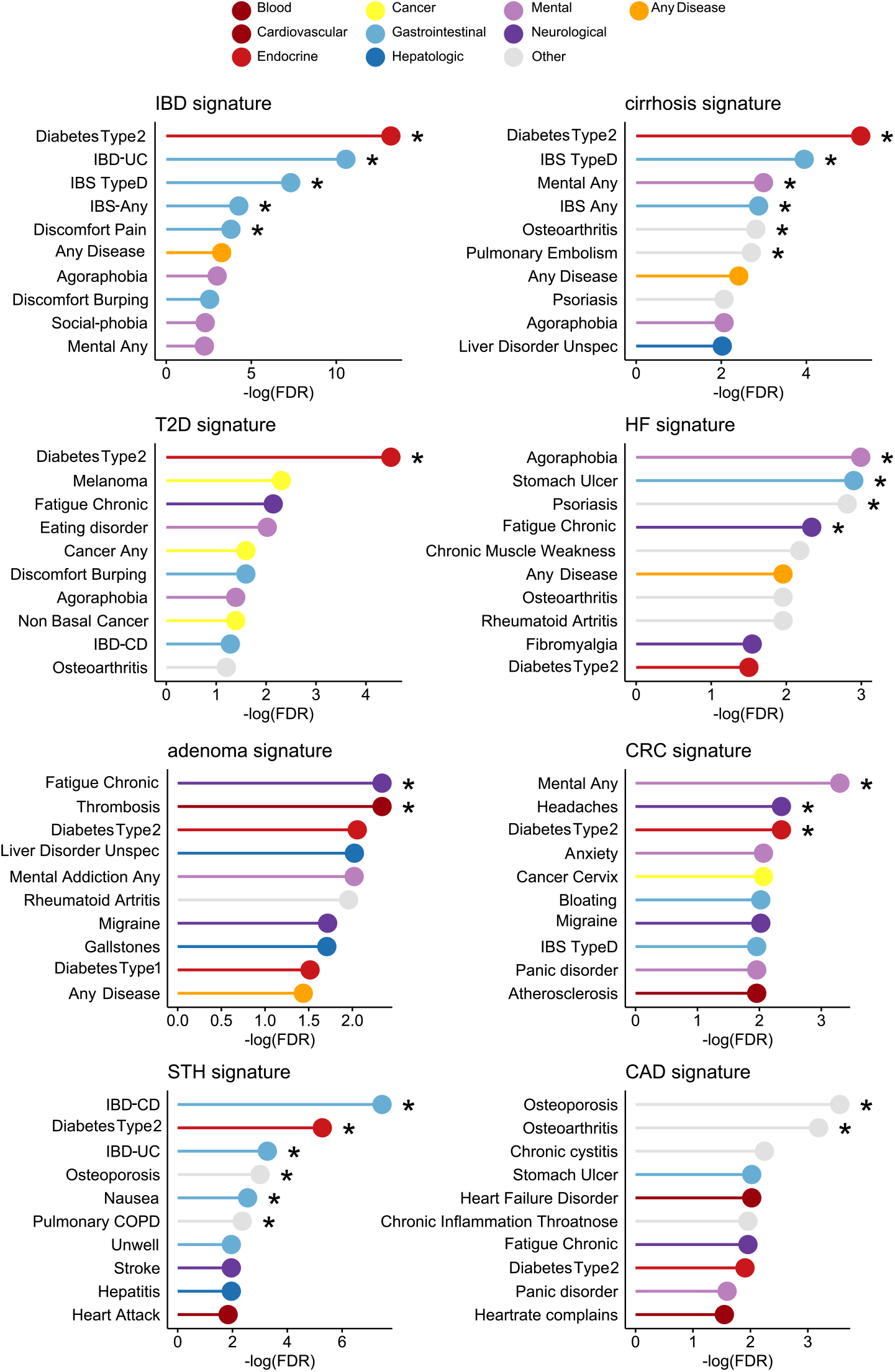
Overrepresented diseases among subjects with high microbial scores. Enrichment analysis was calculated stratifying the population based on the 90 percentile for IBD, cirrhosis, T2D and HF, adenoma, CRC, STH and CAD signature scores. Plots show the top 10 diseases for each signature, with their respective significance, −log(FDR) in the horizontal axis. (*) is for FDR ≤ 0.1. Abbreviations: IBD (intestinal bowel disease), T2D (type 2 diabetes), HF (heart failure), STH soil transmitted Helminths), IBD-UC (ulcerative colitis), CAD (coronary artery disease), IBS (intestinal bowel syndrome), COPD (Chronic obstructive pulmonary disease)

Subjects presenting large values of IBD signature were overrepresented by ulcerative colitis, intestinal bowel syndrome and gastrointestinal discomforts phenotypes, like pain and burping. Another strong signal, cirrhosis, presented hepatic disorders (e.g., hepatitis and gallstones) within the ten most enriched phenotypes, although without statistical significance. Type 2 diabetes was highly enriched in subjects with large values of T2D signature, being the only phenotype significantly enriched in that signature. Given the impact of STH infection in the gastrointestinal tract, it was not unexpected that the signature was enriched in intestinal symptoms like nausea and discomfort, or other gastrointestinal diseases as Crohn’s disease and ulcerative colitis. HF and CAD signatures showed an enrichment in bone disorders like osteoporosis and osteoarthritis, which is in line with the positive reported associations between these bone disorders and cardiovascular problems, all highly prevalent among elderly population [18]. CAD signature was also enriched in heart rate complains and heart failure disorders among the top ten phenotypes. Cervix cancer appeared to be enriched in the CRC signature, although without enough significance.

Finally, the association between significant bacterial signatures and blood measurements was determined using linear regressions and adjusting for covariates (age, sex, BMI, P/B and α-diversity), **Fig. 6**. Several blood measurements resulted to be positively associated with most signatures: triglycerides, glucose and potassium, specific immune cell like neutrophiles, monocytes, lymphocytes, and thrombocytes, as well as HbA1c, pulse rate and systolic pressure. High levels of these indicators are risk factors of several diseases. On the other hand, HDL cholesterol presented negative associations with many signatures.

**Fig 6.**
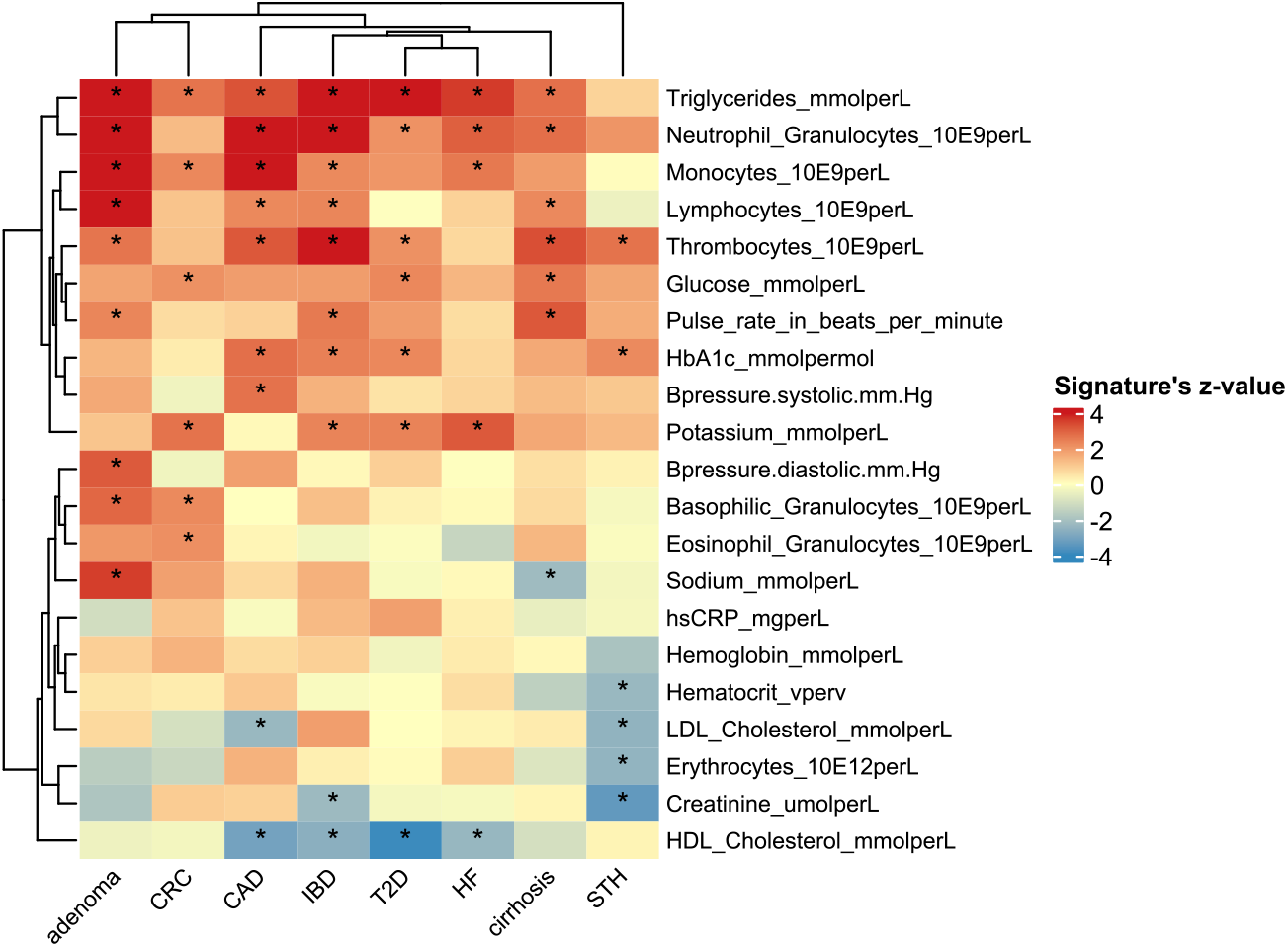
Associations between bacterial signatures and blood measurements. Heatmap colours represent signature’s z-value in the regression adjusted for covariates. Significance is determined by (*) FDR ≤ 0.1

## Discussion

Several attempts have been made to use microbiome as a potential tool for disease diagnostic or prognostic. Instead of a diagnostic tool, our goal was to consider multiple bacterial compositions corresponding to specific physiological processes altered during disease and proof their association with a loss of health status in the general population. We are aware of the difficulties in defining what constitutes a healthy microbiome, if it is even possible. However, we believe it is feasible to identify departures from a healthy state by identifying microbiome patterns associated with imbalances (dysbiosis) or dysfunctions. Doing so, we addressed the principal hypothesis of this study that a certain degree of dysbiosis might already be present before the manifestation of disease.

Two principal steps were performed to address this research. In the first discovery stage, we characterized 16 disease-specific bacterial signatures from publicly available data as a way of measuring microbiome composition under disease. Next, in the validation stage, we quantified the disease-specific bacterial signatures in a general population cohort and tested their association with general health. Results corroborated our initial hypothesis: for most of the signatures, a certain degree of dysbiosis was present without the manifestation of any disease and it was associated with poorer individual’s self-reported general health. Even when adjusting for possible confounders (age, sex, BMI, P/B and α-diversity), some signatures kept significant. This suggests that microbial composition captures certain physiological alterations that might be present before the manifestation of the disease and highlights the potential of microbiome-based models in preventive medicine. In this case, we identified positive associations between 11 disease-specific signatures and general health, from which 6 were further confirmed after adjustment.

Most of the signatures presented significant association but their additional contribution to health prediction given basal covariates and α-diversity was small. A few microbial signatures (IBD, IBD-UC, cirrhosis and HF) provided significant, though modest, improvement in prediction compared to basal model. It is a general trend in microbiome studies, especially related to health and disease, that associations are clearly identified, but high predictions are hard to obtain. Other microbiome-health indices have been proposed that combine the relative abundances of specific microbial species in a univariate score [9, 10, 19, 20]. Though, all of them are intended to measure gut microbiome health, they have been validated according to their accuracy for discriminating between diseased and controls. It is not clear how these indices would perform for assessing general health in non-diseased individuals. This confirms the potential use of microbiome as an additional tool for evaluating health that provides hints of the level of health, but not as a diagnostic tool. On the other hand, alpha diversity is known to be a good measure of microbiome status, given that a higher diversity is usually associated with better health outcomes [21]. Our results corroborate the association between α-diversity and better health and show good performance of chao1 metrics in predicting general health in our cohort.

Although results of our study are promising, we must not overlook its limitations and challenges. The first limitation regards the discovery stage that required combining data from different studies. Though different attempts were followed to reduce batch effects due to study source, we could not always guarantee their entirely removal. A second issue is the robustness of our results under different filtering criteria since a broader range of studies and samples could be considered under less restrictive filters. Third, the model was performed at the species level from shotgun microbiome data processed with Metaphlan3, which was available at the time of the analyses. Probably coming updates of taxonomy assignment, functional profiling, or the integration with of other levels of omics (e.g., genomics, metabolomics or transcriptomics) would provide a more detailed characterization of the analysed diseases. Although these analyses were beyond the scope of this study, they could represent potential future research lines that complement our taxonomic analysis. Moreover, as public microbiome data increases it will be possible to enlarge the number of health conditions and provide a broader view of microbial dysbiosis. Another aspect that we did not address in this work is the geographic specificity of the model. The model was built with data from different geographic areas and validated in the general population of the northern Netherlands. Indeed, the signatures could be ready for additional calibration with geographic-specific microbiome data. An important challenge of our model is its biological interpretation. A more in-depth investigation, perhaps with microbiome functional analysis, would be necessary to associate the different signatures to specific physiological processes.

## Conclusions

In this study, we provide evidence that some dysbiotic microbiome profiles, initially associated with disease, are also present in the general population and correlate with general health. These findings highlight the relevance and potential of microbial information as a tool for preventive medicine and early health assessment. Some of the identified dysbiotic signatures could be used to assess health status in the general population and may serve as a foundation for developing a multidimensional model for gut microbial health evaluation, although the specific mechanisms underlying such associations require further investigation.

## Data Availability

All data produced in the present study are available upon reasonable request to the authors

## Statements & Declarations

### Authors’ contributions

M.P. and M.C. contributed to the study conception and design, execution of analyses, interpretation of results, and preparation and writing the manuscript,. A.K. and A.Z. participated in the validation stage of the analyses and interpretation of results. R.K.W, A.K., A.Z. and J.F. were involved in the data collection. All authors read and approved the final manuscript.

### Ethics approval

This study used publicly available data from previously published studies and open-access databases, for which no additional ethical approval was required.

In addition, data from the LifeLines Cohort Study was used under the cohort’s data access agreement. The LifeLines study was approved by the Medical Ethics Committee of the University Medical Center Groningen (METc UMCG), The Netherlands, and was conducted in accordance with the Declaration of Helsinki. All participants provided written informed consent.

No new data was collected from human participants for the purpose of this study.

### Data availability

The raw microbiome sequencing data, processed microbiome data (including taxonomy, pathway, virulence factor and antibiotic resistance gene profiles) and basic phenotypes (including age, sex and BMI) used in this study are available at the European Genome-Phenome Archive under accession EGAS00001005027. These datasets can be accessed from https://forms.gle/eHeBdXJMXbVvCJRc8 or by email from R.K.W. at the address listed at the EGA data access committee EGAC00001001996. The phenotype data can be requested, for a fee, by filling the application form at https://www.lifelines.nl/researcher/how-to-apply/apply-here.

## References

1. Evans AS. Causation and disease: the Henle-Koch postulates revisited. Yale J Biol Med. 1976;49(2):175–95.

2. Sun Z, Huang S, Zhu P, et al. A microbiome-based index for assessing skin health and treatment effects for atopic dermatitis in children. mSystems. 2019;4:e00293–19. 10.1128/msystems.00293-19

3. Xia GH, You C, Gao XX, et al. Stroke dysbiosis index (SDI) in gut microbiome are associated with brain injury and prognosis of stroke. Front Neurol. 2019;10:397. 10.3389/fneur.2019.00397

4. Beck D, Foster JA. Machine learning classifiers provide insight into the relationship between microbial communities and bacterial vaginosis. BioData Min. 2015;8:1–9. 10.1186/s13040-015-0055-3

5. Wu X, Tang Z, Zhao R, Wang Y, Wang X, Liu S, Zou H. Taxonomic and functional profiling of fecal metagenomes for the early detection of colorectal cancer. Front Oncol. 2023;13:1218056. 10.3389/fonc.2023.1218056

6. Qin J, Li R, Raes J, et al. A human gut microbial gene catalogue established by metagenomic sequencing. Nature. 2010;464:59–65. 10.1038/nature08821

7. The Human Microbiome Project Consortium. Structure, function and diversity of the healthy human microbiome. Nature. 2012;486:207–214. 10.1038/nature11234

8. Salosensaari A, Laitinen V, Havulinna AS, et al. Taxonomic signatures of cause-specific mortality risk in human gut microbiome. Nat Commun. 2021;12:1–8. 10.1038/s41467-021-22962-y

9. Gupta VK, Kim M, Bakshi U, et al. A predictive index for health status using species-level gut microbiome profiling. Nat Commun. 2020;11:18476. 10.1038/s41467-020-18476-8

10. Gacesa R, Kurilshikov A, Vich Vila A, et al. Environmental factors shaping the gut microbiome in a Dutch population. Nature. 2022;604:732–739. 10.1038/s41586-022-04567-7

11. Pasolli E, Schiffer L, Manghi P, et al. Accessible, curated metagenomic data through ExperimentHub. Nat Methods. 2017;14:1023–1024. 10.1038/nmeth.4468

12. Calle ML, Pujolassos M, Susin A. coda4microbiome: compositional data analysis for microbiome cross-sectional and longitudinal studies. BMC Bioinformatics. 2023;24:82. 10.1186/S12859-023-05205-3

13. De Filippo C, Cavalieri D, Di Paola M, et al. Impact of diet in shaping gut microbiota revealed by a comparative study in children from Europe and rural Africa. Proc Natl Acad Sci U S A. 2010;107:14691–14696. 10.1073/pnas.1005963107

14. Roager HM, Licht TR, Poulsen SK, et al. Microbial enterotypes, inferred by the Prevotella-to-Bacteroides ratio, remained stable during a 6-month randomized controlled diet intervention with the new nordic diet. Appl Environ Microbiol. 2014;80:1142–1149. 10.1128/AEM.03549-13

15. Gorvitovskaia A, Holmes SP, Huse SM. Interpreting Prevotella and Bacteroides as biomarkers of diet and lifestyle. Microbiome. 2016;4:1–12. 10.1186/s40168-016-0160-7

16. R Core Team. R: A language and environment for statistical computing. Vienna, Austria: R Foundation for Statistical Computing; 2022.

17. Johnson RA, Wichern DW. Applied multivariate statistical analysis. 6th ed. Upper Saddle River, NJ: Pearson Prentice Hall; 2007.

18. Loncar G, Cvetinovic N, Lainscak M, et al (2020) Bone in heart failure. J Cachexia Sarcopenia Muscle. 2020;11(2):381–393. 10.1002/jcsm.12516

19. Chang D, Gupta VK, Hur B, et al. Gut Microbiome Wellness Index 2 for enhanced health status prediction from gut microbiome taxonomic profiles. bioRxiv. 2023. 10.1101/2023.09.30.560294

20. Zhu J, Xie H, Yang Z, et al. Statistical modeling of gut microbiota for personalized health status monitoring. Microbiome. 2023;11:184. 10.1186/s40168-023-01614-x

21. Behrouzi A, Nafari AH, Siadat SD. The significance of microbiome in personalized medicine.Clin Transl Med. 2019;8(1):16. 10.1186/s40169-019-0232-y

