## Supplementary Material for "Compositional microbiome-based signatures associate with general health status: findings from a large population-based cohort study"

### Disease-specific bacterial signature discovery

#### I. Disease-specific bacterial signatures characterization with *coda4microbiome*

The discovery stage of disease-specific bacterial signatures relies on the data available at *curatedMetagenomicData* (Pasolli et al. 2017), a project that collects metagenomic data from publicly available studies investigating human health and disease. The database contains microbial abundance tables of published studies that have been processed uniformly using Metaphlan3, as described in (Pasolli et al. 2017). Their respective clinical and anthropomorphic data is manually curated by the authors of the database. *curatedMetagenomicData* (version 3.6.2) includes a total of 22.588 samples, from which 21.030 correspond to stool samples from 86 studies investigating up to 51 different health conditions. Disease-specific bacterial signatures characterization was performed on relative abundances at the species taxonomic level. We removed unassigned OTUs (for all taxonomic levels) and filtered metagenomic data based on taxa's prevalence and abundance, including only taxa present in at least 5% of the samples with a mean relative abundance of 0.01 or more. Our analyses consider subjects suffering from specific health conditions (cases) compared to healthy individuals (controls). Stool samples of subjects reporting a given study condition (*e.g.*, T2D) in *curatedMetagenomicData* were labelled as "cases" in our study. Moreover, subjects suffering other co-morbidities, apart from the studied health condition, were also included as "cases" in our study to capture all the variability for that given condition. The control group in every studied condition was composed of individuals from the corresponding selected studies categorized as "healthy" in *curatedMetagenomicData*. Further filters concerning metadata were applied to samples: we included samples with information for possible confounding variables (i.e., BMI, sex, age, diet, and study source) and corrected for them in our analyses to avoid possible bias. Due to a lack of exact ages in many studies, we categorized samples by age groups (newborn, child, school age, adult, senior) instead of filtering by specific years to avoid data loss. Newborns and children were excluded because of high variability in microbiome composition during early life. We also discarded subjects that reported to be in current antibiotic treatment and repeated longitudinal measurements, keeping only the first sample collected. We finally included studies that, after applying the filters, remained with at least 10 healthy and 10 diseased subjects. Overall, we analysed a total of 6.140 subjects, from which 2.852 suffered from 20 different diseases and 3.288 were healthy subjects. **Table S 1** provides detailed information on sample sizes for every health condition.

We adopted the compositional data analysis (CoDA) approach of *coda4microbiome* algorithm (Calle et al. 2023) to address the compositionality of the data. The algorithm analyses relative abundances of some species in relation to other species within the same sample. Specifically, *coda4microbiome* algorithm considers the relative abundances between all pairs of taxa's abundance (log-ratios) and performs an elastic-net penalized regression to select those that are most associated with the outcome variable. Given a phenotype $Y = (Y_1, \ldots, Y_n)$ for *n* subjects and the composition of *K* microbial species for subject *i* by $X_i = (X_{i1}, X_{i2}, \ldots, X_{iK})$, the algorithm considers a generalized linear model containing all possible pairs of log-ratio abundances:

$$g\big(E(Y)\big) = \beta_0 + \sum_{1 \le j < k \ge K} \beta_{jk} \cdot \log\left(X_j / X_k\right) \qquad \text{(Eq. 1)}$$

The regression coefficients in Eq. 1 are estimated to minimize a loss function subject to an elastic-net penalization term on the regression coefficients. The result of the penalized optimization provides a set of selected pairs of microbial species with non-zero coefficients. The linearity of the logarithms allows to express the final signature in terms of single features instead of log-ratios that are difficult to interpret:

$$M = \sum_{1 \leq j < k \geq K} \hat{\beta}_{jk} \cdot \log\left(X_j / X_k\right) = \sum_{j=1}^{K} \hat{\theta}_j \cdot \log\left(X_j\right) \qquad \text{(Eq. 2)}$$

The final signature (Eq. 2) corresponds to a log-contrast function where $\sum_{j=1}^{K} \hat{\theta}_j = 0$, which ensures the scale invariance principal of CoDA and improves interpretability of results. Moreover, the algorithm allows the adjustment for other non-compositional covariates by including them as covariates in the regression model.

*coda4microbiome* allowed to identify a total of 20 disease-specific bacterial signatures (*i.e.*, combination of bacteria that best predicts the disease status compared to healthy controls). Since microbiome is known to be highly influenced by several physiological and environmental factors, we considered sex, age, BMI, the ratio Prevotella/Bacteroides (as a proxy for diet) and study source as possible confounders in our study. In our analysis, all the models were adjusted for these variables when available. Therefore, the predictive models obtained with *coda4microbiome* (referred to c4m) contain both, the microbial signature (M) and the adjustment covariates (B):

$$Y \sim M + B$$

where $Y$ is the outcome variable (*i.e.* disease status): inflammatory bowel disease (IBD), Crohn's disease (IBD-CD), ulcerative colitis (IBD-UC), atherosclerotic cardiovascular disease (ACVD), heart failure (HF), impaired glucose tolerance (IGT), coronary artery disease (CAD), Behcet's disease (BD), pre-hypertension and hypertension, cirrhosis, adenoma and colorectal cancer (CRC), schizophrenia, migraine, Parkinson's disease (PD), asthma and soil transmitted helminths (STH) infection.

To evaluate the actual contribution of the microbiome part (M) in predicting health conditions, we compared, in terms of prediction accuracy, the microbiome-based model c4m ($Y \sim M + B$) with the basal model that only includes the adjustment covariates ($Y \sim B$).

*Table S 1 Datasets used for the characterization of different health conditions*

| Dataset ID | Health condition studied | Co-morbidities in cases | Total N (% cases) | Original study |
|---|---|---|---|---|
| cirrhosis | cirrhosis | hepatitis, ascites, schistose-miasi, wilson's disease | 236 (0.52) | (Qin et al. 2014) |
| CAD | coronary artery disease | T2D | 173 (0.53) | (Vieira-Silva et al. 2020) |
| HF | heart failure | CAD, T2D | 93 (0.13) | (Vieira-Silva et al. 2020) |
| IGT | impaired glucose tolerance | MS | 279 (0.56) | (Karlsson et al. 2013; Vieira-Silva et al. 2020) |
| T2D | type 2 diabetes | none | 830 (0.64) | QinJ_2012, MetaCardis_2020_a, KarlssonFH_2013 |
| ACVD | atheroscle-rotic cardiovas-cular disease | none | 380 (0.55) | (Jie et al. 2017) |
| BD | Beçet's disease | none | 62 (0.32) | (Ye et al. 2018) |
| pre-hypertension | pre-hypertension | none | 97 (0.58) | (Li et al. 2017) |
| hypertension | hypertension | none | 140 (0.71) | (Li et al. 2017) |
| CRC | colorectal cancer | fatty liver, hypertension, hypercholesterolemia, T2D, metastases, cholesterol-lemia | 1319 (0.53) | (Zeller et al. 2014; Feng et al. 2015; Vogtmann et al. 2016; Yu et al. 2017; Hannigan et al. 2018; Dhakan et al. 2019; Thomas et al. 2019; Wirbel et al. 2019; Yachida et al. 2019; Gupta et al. 2019) |
| adenoma | adenoma | fatty liver, hypertension, hypercholeste-rolemia, metastases | 574 (0.36) | (Zeller et al. 2014; Feng et al. 2015; Hannigan et al. 2018; Thomas et al. 2019; Yachida et al. 2019) |
| IBD | inflammato-ry bowel disease | perianal fistula | 467 (0.46) | (Li et al. 2014; Nielsen et al. 2014; Lloyd-Price et al. 2019) |
| IBD-UC | ulcerative colitis | none | 338 (0.26) | (Nielsen et al. 2014; Lloyd-Price et al. 2019) |
| IBD-CD | Crohn's disease | none | 292 (0.14) | (Nielsen et al. 2014; Lloyd-Price et al. 2019) |

| | | | | |
|---|---|---|---|---|
| STH | soil transmitted helminths | none | 153 (0.50) | (Rubel et al. 2020) |
| ME-CFS | myalgic encephalo-myelitis/ chronic fatigue syndrome | none | 100 (0.50) | (Nagy-Szakal et al. 2017) |
| schizophrenia | schizo-phrenia | none | 154 (0.49) | (Zhu et al. 2020) |
| PD | Parkinson disease | none | 42 (0.55) | (Bedarf et al. 2017) |
| migraine | migraine | asthma, generic diabetes | 218 (0.22) | (Xie et al. 2016) |
| asthma | asthma | none | 193 (0.12) | (Xie et al. 2016) |

### II. Overview of all disease-specific bacterial signatures

#### *Signatures characteristics*

The number of bacteria included in the different bacterial signatures span from only 2 taxa (asthma) to 36 (schizophrenia). This number depends on the characteristics of microbiome composition in relation to the outcome, rather than in any other factor like the number of initial features, total number of samples, or the proportion of cases analysed (**Table S 2**). In total, the algorithm selected 191 species associated to 20 different diseases. Up to 79 species were selected for contributing positively to diseases and 75 negatively to diseased phenotypes. Most of them were selected in only one signature: 53 positive and 49 negative diseases. Thirty-seven species were selected in both directions for more than one signature (**Fig. S 1**).

Few species were shared between conditions: 18 species were selected at least in 2 different conditions, and 6 species were shared between 3 conditions. *Clostridium sp. CAG:242* and *Erysipelatoclostridium ramosum* were selected in four different conditions. On the other hand, 15 species were selected for contributing only to healthy phenotypes in more than two signatures. They included mostly short chain fatty acids (SCFA) producers, such as *Firmicutes* bacterium CAG:95, several *Roseburia* species (*R. intestinalis, R. faeci,* and CAG:182), *Eubacterium* sp. CAG:38, *Odoribacter spalachnicus*, *Bifidobacterium pseudocatenulatum*, *Anaerostipes hardus*, *Faecalibacterium prusnitzii*, and *Alistipes putredinis*.

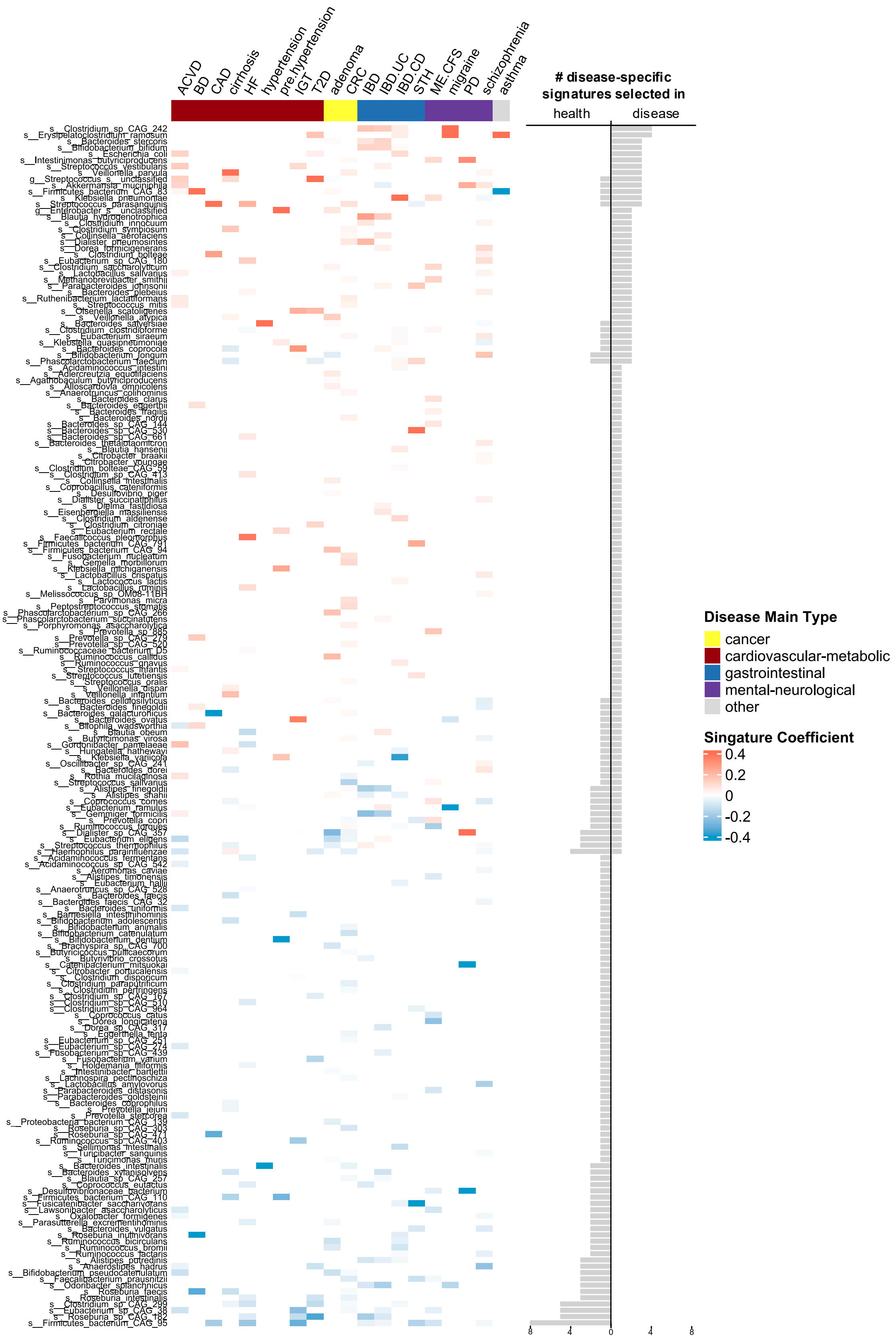


***Fig. S 1*** *Bacterial signatures of the health conditions analysed in the study. The heatmap represents the coefficient of every bacteria within each signature. The two columns on the left show the number of times a bacteria has been selected with positive (red) or negative (blue) coefficient.*

*Prediction power of coda4microbiome models*

Most of the signatures adjusted for sex, age, BMI, P/B and study source, exhibited good predictive accuracy with mean cv-AUCs over 0.7 (**Table S 2**). The highest prediction accuracy was for Crohn's disease (IBD-CD) signature, with a mean cv-AUC (sd) of 0.98 (0.001), followed by cirrhosis 0.95 (0.009). Only four out of twenty signatures presented low prediction power, *i.e.*, mean cv-AUC < 0.7: asthma, migraine, hypertension, and pre-hypertension. We did not identify any correlation between the accuracy of the model and the number of variables included in it, the proportion of cases, or the overall sample size (Pearson correlation coefficient (P-value) of 0.34 (0.14), -0.02 (0.93) and 0.09 (0.71), respectively).

***Table S 2*** *Prediction measurement, mean cv-AUC, of basal models and microbiome-based models by coda4microbiome, c4m models. Differences between means tested with t-test (P-values shown in the last column).*

| Dataset ID | Total samples (% cases) | Number studies | Taxa selected[1] | basal mean cv-AUC (sd) | c4m mean cv-AUC (sd) | |
|---|---|---|---|---|---|---|
| IBD-CD | 292 (14.38) | 2 | 30 | 0.85 (0.048) | 0.98 (0.001) | |
| cirrhosis | 236 (51.69) | 1 | 21 | 0.6 (0.154) | 0.95 (0.009) | *** |
| CAD | 173 (53.18) | 1 | 5 | 0.91 (0,029) | 0.93 (0.018) | · |
| HF | 93 (12.9) | 1 | 22 | 0.72 (0.186) | 0.93 (0.038) | |
| IBD | 467 (45.82) | 3 | 17 | 0.87 (0.042) | 0.91 (0.014) | * |
| IGT | 279 (55.56) | 2 | 13 | 0.87 (0.064) | 0.88 (0.016) | |
| IBD-UC | 338 (26.04) | 2 | 27 | 0.72 (0.125) | 0.87 (0.021) | ** |
| T2D | 830 (64.22) | 3 | 13 | 0.82 (0.027) | 0.87 (0.012) | ** |
| ACVD | 380 (55.26) | 1 | 25 | 0.69 (0.053) | 0.85 (0.028) | *** |
| STH | 153 (50.33) | 1 | 12 | 0.72 (0.158) | 0.84 (0.023) | · |
| ME/CFS | 100 (50) | 1 | 22 | 0.62 (0.170) | 0.83 (0.049) | ** |
| CRC | 1319 (53.15) | 11 | 46 | 0.57 (0.051) | 0.79 (0.008) | *** |
| BD | 62 (32.26) | 1 | 7 | 0.49 (0.111) | 0.78 (0.012) | |
| schizophrenia | 154 (49.35) | 1 | 36 | 0.67 (0.071) | 0.77 (0.041) | ** |
| adenoma | 574 (36.41) | 5 | 29 | 0.73 (0.063) | 075 (0.026) | |
| PD | 42 (54.76) | 1 | 5 | 0.61 (0.120) | 0.75 (0.077) | |
| pre-hypertension | 97 (57.73) | 1 | 8 | 0.56 (0.087) | 0.66 (0.078) | |
| migraine | 218 (22.48) | 1 | 5 | 0.58 (0.087) | 0.65 (0.051) | |
| asthma | 193 (12.44) | 1 | 2 | 0.51 (0.042) | 0.60 (0.058) | |
| hypertension | 140 (70.71) | 1 | 2 | 0.58 (0.039) | 0.60 (0.017) | |

T-test between mean cv-AUCs significance: *** P-value ≤ 0.0001; ** P-value ≤ 0.01; * P-value ≤ 0.05

[1]Only applied to c4m models

We assessed the additional contribution of microbiome composition in predicting the presence or absence of disease by comparing c4m models with basal models in terms of their mean cv-AUC. Basal models showed lower predictions compared to c4m models, suggesting that the addition of microbiome information improved prediction. Particularly, eight basal models improved significatively their prediction when considering microbiome information: cirrhosis, IBD, IBD-UC, T2D, ACVD, ME/CFS, CRC, and schizophrenia. In some cases, basal models already performed very good, with cv-AUC above 0.7, and although the addition of microbiome increased prediction, it was not significant (IBD-CD, adenoma, IGT, CAD, STH and HF). Some of these diseases are known to be strongly associated to sex, age and BMI, thus the information regarding covariates had high prediction capacity, difficult to improve.

*Accounting for non-compositional covariates*

Adjusting for covariates, like age, BMI, gender, study, and P/B in this case, is a recommended approach to correct for possible confounders. However, the complete removal of covariates' effects from the resulting models is not always guaranteed. To inspect for possible remaining bias due to confounding, even after adjustment, we analysed the interactions of covariates with the predictions and the residuals of the final models. If covariates' effects were removed, no interaction should be observed.

No remaining bias due to variables age and sex was observed in the final models. No signature presented interactions between predictions and age and sex which means that discrimination between samples is driven by real differences in microbiome composition and not to other variables like age or sex. The assessment of models' residuals in relation to sex and age gave concordant results: for most of the models, we did not find significant differences in the residuals within sex and age categories. However, we identified some exceptions: IBD-UC presented differences among adults and senior individuals; and CAD and adenoma between males and females. Regarding P/B and BMI, we did not find important correlations between residuals and these two covariates in our models.

In datasets including more than one study (*e.g.*, IBD, IBD-UC, IBD-CD, adenoma, CRC, T2D and IGT), the influence caused by the source of the data might be considerable when models are not well adjusted. In our analysis, after adjusting by study, all signatures presented significant differences between cases and controls within each study with concordant directions: healthy controls presented lower microbial scores than cases. The analysis of residuals is an additional test that provides more insights about the effect of the study source in our models. We did not find differences in the residuals among studies for most of the studies, except for IBD-CD and IBD models.

### Assessment of disease-specific bacterial signatures in the general population

The assessment of disease-specific bacterial signatures association with general health (GH) in the general population was analysed in three ways: as an ordinal numeric variable (GH), a binary variable (GHbi) grouping "good health" (excellent to good) vs. "bad health" (mediocre and poor), and another binary version (GHbix) including only extreme phenotypes: "excellent" in the good health group and “mediocre” + “poor” in the bad health group. Following the same methodology explained in the article, the following results correspond to analyses performed taking into account either ordinal GH or binary GH (GHbi and GHbix) as outcome variable for general health.

*Table S 3 Kendall trend test results between bacterial signatures scores and general health status (from excellent to poor). Results include Kendall's tau and FDR adjusted P-value.*

| | **Kendall's tau** | **FDR** |
|---|---|---|
| IBD | 0,077 | <0,001 |
| CAD | 0,074 | <0,001 |
| cirrhosis | 0,066 | <0,001 |
| STH | 0,050 | <0,001 |
| IBD.UC | 0,049 | <0,001 |
| T2D | 0,046 | <0,001 |
| HF | 0,041 | <0,001 |
| CRC | 0,037 | <0,001 |
| adenoma | 0,037 | <0,001 |
| PD | 0,027 | 0,011 |
| IBD.CD | 0,026 | 0,009 |
| IGT | 0,020 | 0,050 |
| ME.CFS | 0,013 | 0,201 |
| ACVD | -0,003 | 0,781 |
| schizophrenia | -0,008 | 0,414 |
| BD | -0,026 | 0,009 |

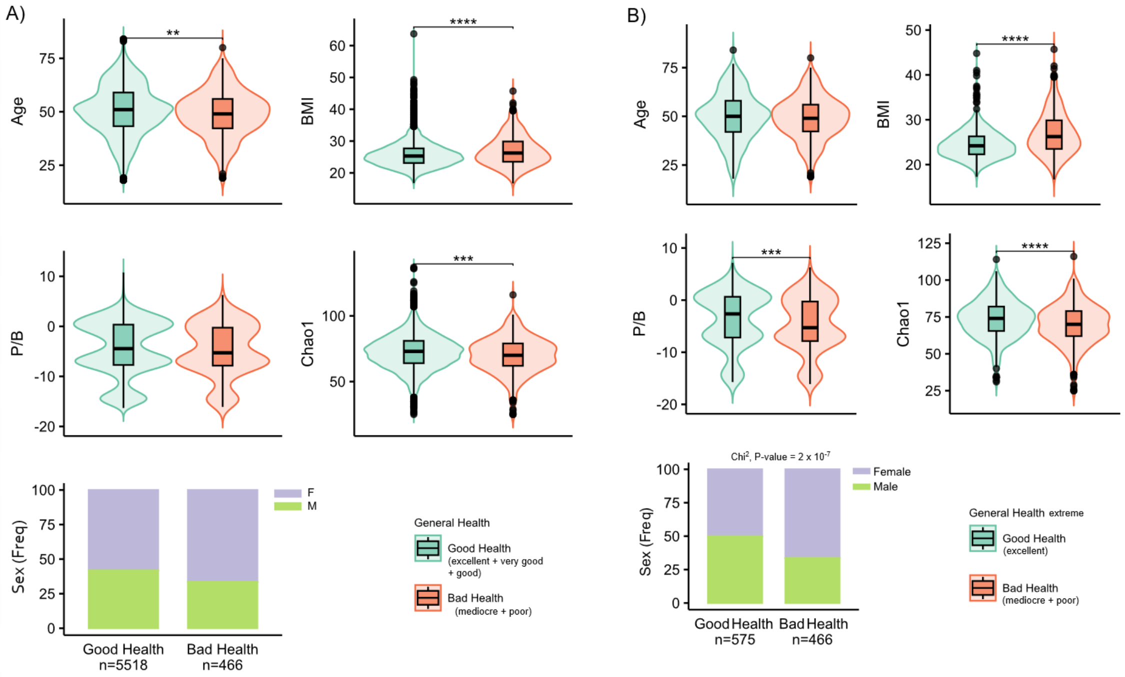


***Fig. S 2** Basal variables distribution across binary categories of General Health. Age, BMI, P/B, chao1 and Sex are represented for (A) categories of GHbi: "Good Health" (excellent + very good + geood) and "Bad Health"(mediocre + poor); and (B) categories of GHbix: "Good Health" (excellent) and "Bad Health"(mediocre + poor). The number of samples within every group is reported (n). Wilcoxon P-value significance: **** ≤ 0.0001, *** ≤ 0.001, ** ≤ 0.01, * ≤ 0.05.*

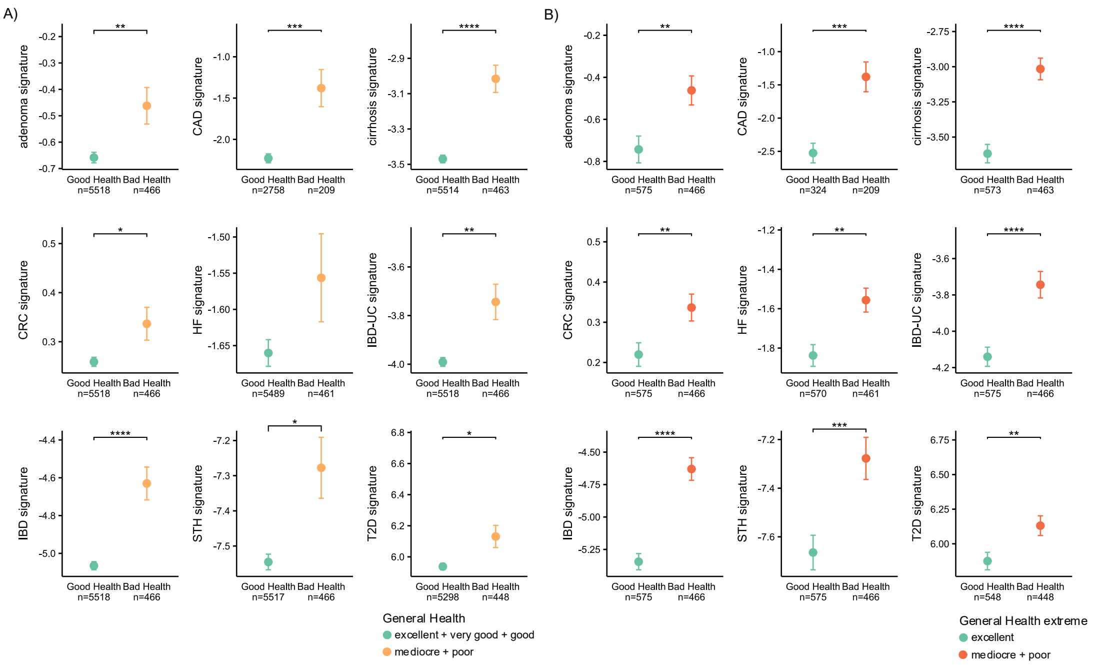


*Fig. S 3 Microbial disease-specific signatures scores of good and bad health. Mean and confidence intervals for adenoma, CAD, cirrhosis, CRC, HF, IBDUC, IBD, STH and T2D according to binary General Health (A) and extreme binary General Health (B) categories.*

GHbi ~ age + sex + BMI + P/B + α-div + M

| | | |
|---|---|---|
| 4.478 (8e-06) | * | cirrhosis |
| 4.083 (4e-05) | * | CAD |
| 3.468 (5e-04) | * | IBD |
| 2.924 (0.003) | * | IBD-UC |
| 2.262 (0.02) | * | adenoma |
| 2.024 (0.04) | * | STH |
| 1.262 (0.2) | | T2D |
| 1.13 (0.3) | | CRC |
| 0.8 (0.4) | | HF |
| 0.182 (0.9) | | IBD-CD |
| 0.178 (0.9) | | IGT |
| 0.102 (0.9) | | BD |
| -0.441 (0.7) | | PD |
| -0.739 (0.5) | | schizophrenia |
| -0.869 (0.4) | | ACVD |
| -1.862 (0.06) | | ME/CFS |

GHbix ~ age + sex + BMI + P/B + α-div + M

| | | |
|---|---|---|
| 4.562 (5e-06) | * | IBD |
| 4.102 (4e-05) | * | CAD |
| 3.699 (2e-04) | * | IBD-UC |
| 3.444 (6e-04) | * | cirrhosis |
| 2.7 (0.007) | * | HF |
| 1.628 (0.1) | | adenoma |
| 1.425 (0.2) | | T2D |
| 0.962 (0.3) | | STH |
| 0.94 (0.3) | | CRC |
| 0.864 (0.4) | | IGT |
| 0.755 (0.5) | | PD |
| 0.455 (0.6) | | schizophrenia |
| -0.224 (0.8) | | IBD-CD |
| -1.034 (0.3) | | ACVD |
| -1.046 (0.3) | | BD |
| -1.751 (0.08) | | ME/CFS |

**Signature's z-value (p-value)**

-10 -5 0 5 10

**ANOVA significance**
(basal models vs M model)
* P-value < 0.05

***Fig. S 4*** *Microbial signatures association with general health GHbi and GHbix. Heatmap colours represent t-values of every signature, M, (P-values in brackets). Positive values are for signature associated with poorer general health, while negative values indicate association with better general health. The analyses of variance between each M model and their respective basal model were performed with ANOVA, and its statistical significance is indicated with (*).*

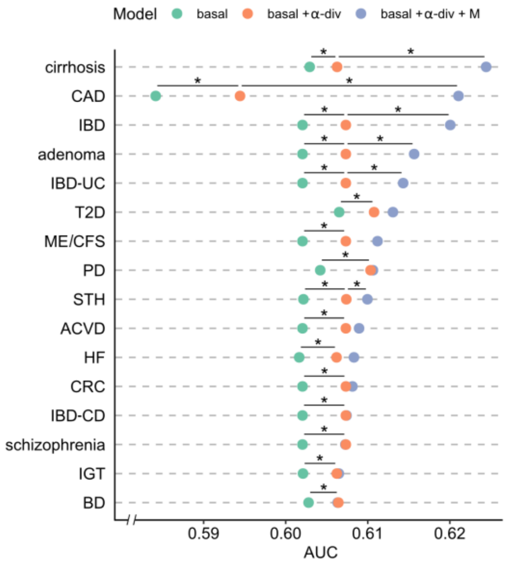


***Fig. S 5*** *Contribution of microbiome in predicting general health GHbi. The prediction performance in terms AUC of basal models, models with α-div, and models with α-div + bacterial signatures (M). (*) is for significant improvement between models, assessed with ANOVA, within 95% confidence level.*

*Table S 4 Contribution of microbiome in GHnum regression models. ANOVA for nested model statistical significance is reported by P-values.*

| Signatures (M) | ANOVA P-value | |
|---|---|---|
| | basal *vs* basal + α-div | basal + α-div *vs* basal + α-div + M |
| IBD | **5,22 x 10-7** | **9,38 x 10-8** |
| CAD | **2,28 x 10-3** | **6,61 x 10-7** |
| IBD-UC | **5,22 x 10-7** | **1,07 x 10-5** |
| cirrhosis | **6,24 x 10-7** | **3,84 x 10-4** |
| HF | **4,98 x 10-7** | **9,37 x 10-3** |
| BD | **1,04 x 10-6** | **1,42 x 10-2** |
| adenoma | **5,22 x 10-7** | **2,29 x 10-2** |
| STH | **5,36 x 10-7** | 5,17 x 10-2 |
| T2D | **1,92 x 10-7** | 5,89 x 10-2 |
| CRC | **5,22 x 10-7** | 1,22 x 10-1 |
| PD | **1,68 x 10-6** | 2,23 x 10-1 |
| ACVD | **5,22 x 10-7** | 3,00 x 10-1 |
| ME/CFS | **5,22 x 10-7** | 3,52 x 10-1 |
| schizophrenia | **5,22 x 10-7** | 6,52 x 10-1 |
| IGT | **1,09 x 10-6** | 7,74 x 10-1 |
| IBD-CD | **5,22 x 10-7** | 9,26 x 10-1 |

### References


Bedarf JR, Hildebrand F, Coelho LP, et al (2017) Functional implications of microbial and viral gut metagenome changes in early stage L-DOPA-naïve Parkinson's disease patients. Genome Med 9:. https://doi.org/10.1186/s13073-017-0428-y

Calle ML, Pujolassos M, Susin A (2023) coda4microbiome: compositional data analysis for microbiome cross-sectional and longitudinal studies. BMC Bioinformatics 24:. https://doi.org/10.1186/S12859-023-05205-3

Dhakan DB, Maji A, Sharma AK, et al (2019) The unique composition of Indian gut microbiome, gene catalogue, and associated fecal metabolome deciphered using multi-omics approaches. Gigascience 8:. https://doi.org/10.1093/gigascience/giz004

Feng Q, Liang S, Jia H, et al (2015) Gut microbiome development along the colorectal adenoma-carcinoma sequence. Nat Commun 6:. https://doi.org/10.1038/ncomms7528

Gupta A, Dhakan DB, Maji A, et al (2019) Association of Flavonifractor plautii, a Flavonoid-Degrading Bacterium, with the Gut Microbiome of Colorectal Cancer Patients in India. mSystems 4:. https://doi.org/10.1128/msystems.00438-19

Hannigan GD, Duhaime MB, Ruffin MT, et al (2018) Diagnostic Potential and Interactive Dynamics of the Colorectal Cancer Virome. American Society for Microbiology 9:e02248-18. https://doi.org/10.1128/mBio

Jie Z, Xia H, Zhong SL, et al (2017) The gut microbiome in atherosclerotic cardiovascular disease. Nat Commun 8:. https://doi.org/10.1038/s41467-017-00900-1

Karlsson FH, Tremaroli V, Nookaew I, et al (2013) Gut metagenome in European women with normal, impaired and diabetic glucose control. Nature 498:99–103. https://doi.org/10.1038/nature12198

Li J, Jia H, Cai X, et al (2014) An integrated catalog of reference genes in the human gut microbiome. Nat Biotechnol 32:834–841. https://doi.org/10.1038/nbt.2942

Li J, Zhao F, Wang Y, et al (2017) Gut microbiota dysbiosis contributes to the development of hypertension. Microbiome 5:. https://doi.org/10.1186/s40168-016-0222-x

Lloyd-Price J, Arze C, Ananthakrishnan AN, et al (2019) Multi-omics of the gut microbial ecosystem in inflammatory bowel diseases. Nature 569:655–662. https://doi.org/10.1038/s41586-019-1237-9

Nagy-Szakal D, Williams BL, Mishra N, et al (2017) Fecal metagenomic profiles in subgroups of patients with myalgic encephalomyelitis/chronic fatigue syndrome. Microbiome 5:44. https://doi.org/10.1186/S40168-017-0261-Y

Nielsen HB, Almeida M, Juncker AS, et al (2014) Identification and assembly of genomes and genetic elements in complex metagenomic samples without using reference genomes. Nat Biotechnol 32:822–828. https://doi.org/10.1038/nbt.2939

Pasolli E, Schiffer L, Manghi P, et al (2017) Accessible, curated metagenomic data through ExperimentHub. Nat Methods 14:1023–1024. https://doi.org/10.1038/nmeth.4468

Qin N, Yang F, Li A, et al (2014) Alterations of the human gut microbiome in liver cirrhosis. Nature 513:59–64. https://doi.org/10.1038/nature13568

Rubel MA, Abbas A, Taylor LJ, et al (2020) Lifestyle and the presence of helminths is associated with gut microbiome composition in Cameroonians. Genome Biol 21:. https://doi.org/10.1186/s13059-020-02020-4

Thomas AM, Manghi P, Asnicar F, et al (2019) Metagenomic analysis of colorectal cancer datasets identifies cross-cohort microbial diagnostic signatures and a link with choline degradation. Nat Med 25:667–678. https://doi.org/10.1038/s41591-019-0405-7

Vieira-Silva S, Falony G, Belda E, et al (2020) Statin therapy is associated with lower prevalence of gut microbiota dysbiosis. Nature 581:310–315. https://doi.org/10.1038/s41586-020-2269-x

Vogtmann E, Hua X, Zeller G, et al (2016) Colorectal cancer and the human gut microbiome: Reproducibility with whole-genome shotgun sequencing. PLoS One 11:. https://doi.org/10.1371/journal.pone.0155362

Wirbel J, Pyl PT, Kartal E, et al (2019) Meta-analysis of fecal metagenomes reveals global microbial signatures that are specific for colorectal cancer. Nat Med 25:679–689. https://doi.org/10.1038/s41591-019-0406-6

Xie H, Guo R, Zhong H, et al (2016) Shotgun Metagenomics of 250 Adult Twins Reveals Genetic and Environmental Impacts on the Gut Microbiome. Cell Syst 3:572-584.e3. https://doi.org/10.1016/j.cels.2016.10.004

Yachida S, Mizutani S, Shiroma H, et al (2019) Metagenomic and metabolomic analyses reveal distinct stage-specific phenotypes of the gut microbiota in colorectal cancer. Nat Med 25:968–976. https://doi.org/10.1038/s41591-019-0458-7

Ye Z, Zhang N, Wu C, et al (2018) A metagenomic study of the gut microbiome in Behcet's disease. Microbiome 6:. https://doi.org/10.1186/s40168-018-0520-6

Yu J, Feng Q, Wong SH, et al (2017) Metagenomic analysis of faecal microbiome as a tool towards targeted non-invasive biomarkers for colorectal cancer. Gut 66:70–78. https://doi.org/10.1136/gutjnl-2015-309800

Zeller G, Tap J, Voigt AY, et al (2014) Potential of fecal microbiota for early-stage detection of colorectal cancer. Mol Syst Biol 10:766. https://doi.org/10.15252/MSB.20145645

Zhu F, Ju Y, Wang W, et al (2020) Metagenome-wide association of gut microbiome features for schizophrenia. Nat Commun 11:. https://doi.org/10.1038/s41467-020-15457-9
